# Orthostatic intolerance in adults with long COVID was not associated with postural orthostatic tachycardia syndrome

**DOI:** 10.1101/2021.12.19.21268060

**Authors:** Ann Monaghan, Glenn Jennings, Feng Xue, Lisa Byrne, Eoin Duggan, Roman Romero-Ortuno

## Abstract

In this observational cross-sectional study, we investigated predictors of orthostatic intolerance (OI) in adults with long COVID. Participants underwent a 3-minute active stand (AS) with Finapres® NOVA, followed by a 10-minute unmedicated 70-degree head-up tilt test. 85 participants were included (mean age 46 years, range 25-78; 74% women), of which 56 (66%) reported OI during AS (OI_AS_). OI_AS_ seemed associated with female sex, more fatigue and depressive symptoms, and greater inability to perform activities of daily living (ADL), as well as a higher heart rate (HR) at the lowest systolic blood pressure (SBP) point before the 1^st^ minute post-stand (mean HR_nadir_: 88 vs 75 bpm, P=0.004). In a regression model also including age, sex, fatigue, depression, ADL inability, and peak HR after the nadir SBP, HR_nadir_ was the only OI_AS_ predictor (OR=1.09, 95% CI: 1.01-1.18, P=0.027). 22 participants had initial (iOH) and 5 classical (cOH) orthostatic hypotension, but neither correlated with OI_AS_. 71 participants proceeded to tilt, of which 28 had OI during tilt (OI_tilt_). Of the 53 who had a 10-minute tilt, 7 (13%) fulfilled hemodynamic postural orthostatic tachycardia syndrome (POTS) criteria, but 6 did not report OI_tilt_. OI_AS_ was associated with a higher initial HR on AS, which after 1 minute equalized with the non-OI_AS_ group. Despite these initial orthostatic HR differences, POTS was infrequent and largely asymptomatic. ClinicalTrials.gov Identifier: NCT05027724 (retrospectively registered on August 30, 2021).

## 1 Introduction

Long COVID or post-COVID-19 syndrome first gained recognition among social support groups and later in scientific and medical communities (Yong, 2021). This condition is not well understood as it affects COVID-19 survivors at all ages and levels of disease severity, with or without pre-existing comorbidities, and regardless of hospitalisation status (Vanichkachorn et al., 2021;Yong, 2021). A common symptom is fatigue, with or without organ-specific symptoms (Jennings et al., 2021;Rogers et al., 2021), which may result in negative impacts on resumption of functional and occupational activities (Yan et al., 2021). A systematic review reported that symptoms of mild COVID-19 may persist after 3 weeks in a third of patients (van Kessel et al., 2021). Another study reported that up to one in four patients with mild COVID-19 were still experiencing symptoms after one year (Rank et al., 2021); however, data on the exact prevalence and long-term effects of long COVID are still lacking (Zarei et al., 2021), with an urgent need for research in different populations and settings (Michelen et al., 2021). To aid clinicians and researchers, on 6 October 2021 the World Health Organization (WHO) issued a clinical case definition of post COVID-19 condition, obtained by a Delphi consensus (WHO, 2021), as follows: “Post COVID-19 condition occurs in individuals with a history of probable or confirmed SARS-CoV-2 infection, usually 3 months from the onset of COVID-19 with symptoms that last for at least 2 months and cannot be explained by an alternative diagnosis. Common symptoms include fatigue, shortness of breath, cognitive dysfunction but also others, which generally have an impact on everyday functioning. Symptoms may be new onset, following initial recovery from an acute COVID-19 episode, or persist from the initial illness. Symptoms may also fluctuate or relapse over time”.

The neurological and cardiovascular overlap in some long COVID symptoms, and in particular the reported occurrence of orthostatic intolerance (OI) (Dani et al., 2021;Paterson et al., 2021;Shah et al., 2021), have raised the hypothesis as to whether some long COVID patients could have measurable autonomic nervous system impairments (Del Rio et al., 2020;Goldstein, 2020;Keyhanian et al., 2020;Barizien et al., 2021;Becker, 2021;Larsen et al., 2021) such as orthostatic hypotension (OH) or postural orthostatic tachycardia syndrome (POTS) (Blitshteyn and Whitelaw, 2021;Johansson et al., 2021;Raj et al., 2021). In this light, we conducted a cross-sectional observational study on a cohort of long COVID participants to fulfil the following objectives: (1) establish the prevalence of OI, both during an active stand (AS) test and a tilt test; (2) establish the prevalence of OH and POTS in this cohort; and (3) study haemodynamic and non-haemodynamic predictors of OI.

## 2 Methods

### 2.1 Study and cohort description

This was a cross-sectional observational study on a participant cohort recruited for the TROPIC (Technology assisted solutions for the Recognition of Objective Physiological Indicators of post-Coronavirus-19 fatigue) investigation at Trinity College Dublin and St James’s Hospital Dublin, Ireland. The study received full ethical and regulatory approvals. For the reporting, we followed STROBE guidelines (von Elm et al., 2007).

Participants were eligible for inclusion under all the following criteria: (1) age 18 years or older; (2) history of confirmed or suspected SARS-CoV-2 infection; (3) experiencing prolonged symptoms such as fatigue; (4) able to mobilise independently (with or without aid); (5) able to transfer independently or with minimal assistance of one person from lying to standing; and (6) able to give informed consent.

Participants were recruited from the following sources in our hospital: (1) falls and syncope unit; (2) geriatric day hospital; (3) post COVID-19 outpatient clinic; (4) staff who had contracted COVID-19; and (5) participants from earlier post-COVID-19 research who had consented to be contacted for further studies. In addition, we also considered (6) self-referrals. COVID-19 and non-COVID-19 exclusion criteria for enrolment are outlined in the Supplementary Information (section 1).

Prior to enrolment, participants were provided with a Participant Information Leaflet explaining the aims and procedures of the study. All participants provided explicit, informed, and voluntary consent to partake in the study, were explained the benefits and risks of participating in the research, and had the opportunity to discuss the study and ask questions. Participants were given the opportunity to withdraw from the study at any point and to forego completing components of the assessment protocol as desired.

### 2.2 Procedures

Participants underwent a 3-minute active stand (AS) with Finapres® NOVA, followed by a 10-minute unmedicated 70-degree head-up tilt test. During both, participants had frontal lobe oxygenation monitoring via PortaLite® near-infrared spectroscopy (NIRS). All testing procedures complied with the local hand hygiene, sanitation, personal protective equipment (PPE), and research training protocols. We also considered international best practice recommendations for autonomic testing during the COVID-19 pandemic (Figueroa et al., 2020;Guaraldi et al., 2020;Sinn et al., 2021).

For the active stand, participants underwent a lying-to-standing orthostatic test with non-invasive beat-to-beat blood pressure monitoring using digital photoplethysmography (Finapres® NOVA, Finapres Medical Systems, Amsterdam, The Netherlands). The height correction unit was zeroed and implemented as per manufacturer’s specifications. A 5-lead continuous electrocardiogram (ECG) was acquired throughout the test. During the supine rest period, an oscillometric brachial blood pressure measurement was obtained from the non-monitored (right) arm for calibration purposes, once the PhysioCal repetition rate was 70 beats or more (Wesseling, 1996). After at least 5 minutes of uninterrupted supine rest, a total lying duration of no more than 10 minutes, and a 10-second countdown, participants were asked to stand, unaided, as quick as possible. The PhysioCal was turned off just before the stand and switched back on at 1-minute post-stand. After standing, SBP, DBP, and HR were monitored for three minutes. Throughout the recording, participants were asked to remain motionless and in silence with the monitored arm (left) resting extended by the side, except for reporting any symptoms of concern. Immediately after the stand, and at the end of the test, participants were asked to report whether they had felt any symptoms of dizziness, light-headedness, or any other abnormal symptoms.

For the tilt procedure, which was medically supervised and started after a brief non-monitored break following AS, participants were affixed to an electrically motorized tilt table with footboard support and approximately 10 seconds of travel time between 0° and 70° (Agasan KT-1060/E, AGA Sanitätsartikel GmbH, Löhne, Germany). Throughout the tilt, participants underwent Finapres® NOVA monitoring (with PhysioCal on and continuous ECG monitoring) during an initial period of uninterrupted supine rest of at least 5 minutes (with a total lying duration of no more than 10 minutes) and a subsequent head-up tilt to 70° for 10 minutes or until symptoms developed. An oscillometric brachial blood pressure was also obtained during supine rest. During the head-up tilt phase, participants were asked to report whether they felt any symptoms of dizziness, light-headedness, or any other abnormal symptoms, at which point participants were offered to be tilted down. Even without symptoms, if the head-up tilt elicited hypotension (defined as SBP < 90 mmHg), the tilt was aborted.

For the NIRS-based monitoring of regional cerebral oxygenation of the left frontal lobe during both AS and tilt, we used an optical sensor (PortaLite®, Artinis Medical Systems B.V., Elst, The Netherlands), applied approximately 3 cm to the left of the midline of the forehead and 3.5 cm above the bridge of the nose. A close-woven bandage was affixed around the head over the sensor to remove ambient lighting and to exert comfortable pressure for effective contact between the probe and the skin.

### 2.3 Hemodynamic data extraction

For SBP, DBP and HR, values were noted at the various timepoints of AS and head-up tilt from the Finapres® NOVA display screen in accordance with the following standard operating procedure (SOP): baseline values were collected at 60 seconds prior to AS or head-up tilt, and subsequently at the start of every minute after each procedure. As regards nadir values, for the AS they were noted at the lowest point of SBP following completion of standing and prior to the first minute post-stand; in the case of the tilt, they were noted at the lowest point of SBP reached between completion of the head-up tilt manoeuvre and prior to the first minute post-tilt. For the AS, we also modelled the peak HR after the nadir SBP, defined as the maximum of the HR readings obtained at 1, 2 and 3 minutes. NIRS values were noted following the same SOP from a laptop display connected to the PortaLite® device via OxySoft® software (version 3.2.70), from which we extracted Tissue Saturation Index (TSI) values as the percentage ratio of oxygenated haemoglobin concentration to the total concentration of haemoglobin (Claffey et al., 2020).

### 2.4 Orthostatic hypotension definitions

Initial orthostatic hypotension (iOH) on AS was defined as a difference of >40 mmHg SBP and/or >20 mmHg DBP between baseline and nadir values (Freeman et al., 2011).

Classical orthostatic hypotension on AS (cOH_AS_) was defined as a difference of ≥20 mmHg SBP and/or ≥10 DBP between each baseline value and its minimum reading between minutes 1, 2, and 3 (Freeman et al., 2011). Nadir values were not included in this definition for clear differentiation with iOH and to better reflect cOH_AS_ as normally measured in routine clinical practice with an interval measurement device (Breeuwsma et al., 2018).

Classical orthostatic hypotension on tilt (cOH_tilt_) was defined as a difference of ≥20 mmHg SBP and/or ≥10 DBP between each baseline value and its minimum reading between nadir and minutes 1, 2, and 3. Nadir was included in this case because iOH is only associated with active rising (Wieling et al., 2007).

### 2.5 POTS hemodynamic definition

We computed the maximum HR between nadir and minutes 1 to 10 (or the available minutes in case of early tilt termination), to which we subtracted baseline HR, and POTS was defined in hemodynamic terms as a difference of ≥30 bpm in the absence of cOH_tilt_ (Freeman et al., 2011).

### 2.6 Other measures

For the characterisation of the cohort, we collected measures including:

- Demographics: age, sex.
- Anthropometrics: body mass index (Kg/m^2^).
- Proportion of third level education (i.e., primary university degree or higher).
- Past medical history including previous or current smoker, hypertension, heart disease (e.g., previous heart attack, angina, congestive heart failure, atrial fibrillation), diabetes mellitus (yes or no).
- Current medications including being on an antihypertensive, beta blocker, antidepressant, or benzodiazepine (yes or no).
- COVID-19 history: date of COVID-19 diagnosis; hospitalisation status (at least 1 overnight stay: yes or no); current symptomatology (from a structured questionnaire including 41 possible symptoms: yes or no for each), and interference with activities of daily living (ADL) (“In the past month, I have had too little energy to do the things I wanted to do”: yes or no).
- The 11-item Chalder Fatigue Scale (CFQ), a self-rating scale developed to measure the severity of physical and mental fatigue (Cella and Chalder, 2010). We employed the Likert scoring system, with an overall scale range from 0 (minimum) to 33 (maximum fatigue).
- The 20-item Center for Epidemiological Studies Depression (CES-D) scale (Radloff, 1977). Scores range from 0 to 60, with higher scores indicating greater depressive symptoms.
- The 22-item Impact of Event Scale – Revised (IES-R) (Creamer et al., 2003), which measured post-traumatic stress disorder (PTSD) symptoms in specific relation to participants’ COVID-19 illness (minimum: 0; maximum: 88).
- Five chair stands time as a measure of functional lower extremity strength (Munoz-Bermejo et al., 2021): time (in seconds) it took a participant to transfer as quick as possible from a seated to a standing position and back to sitting five times.

### 2.7 Statistical analyses

Statistics were computed with IBM® SPSS® Statistics for Windows, Version 26.0, Armonk, NY: IBM Corp. Descriptives were given with count and percentage (%), mean with standard deviation (SD), median with interquartile range (IQR), and range. We utilised the SPSS Chart Builder to visualise hemodynamic differences between subgroups via cluster line chart with representation of 95% confidence intervals (CI) around means. To compare characteristics between subgroups, we utilised the non-parametric 2-sided Mann-Whitney U test for continuous variables, and the Chi-square test for dichotomous characteristics. In the latter case, we used the 2-sided Fisher’s exact test when at least one cell had an expected count of <5. To establish independent predictors of dichotomous group membership, we computed logistic regression models, and for each predictor extracted the Odds Ratio (OR) and 95% CI for the OR. Multicollinearity checks were conducted. Statistical significance was defined as P<0.05.

### 2.8 Ethical approval

This study received full approval by the St James’s Hospital/Tallaght University Hospital Joint Research Ethics Committee (Submission Number: 104: TROPIC; Approval Date: 4 May 2021) and the St James’s Hospital Research & Innovation Office (Reference: 6566; Approval Date: 14 May 2021). The study was performed in accordance with the ethical standards laid down in the 1964 Declaration of Helsinki and its later amendments. All participants gave their informed consent prior to their inclusion in the study. All aspects of the study were executed in compliance with the General Data Protection regulation (GDPR), and Irish regulations including the Health Research Regulations and the Data Protection Act 2018.

## 3 Results

Of 92 consecutive participants recruited to the study between May and September 2021, 85 (92.4%) had an AS. Mean age was 46.0 years (SD 10.2, range 25-78), and 63 (74.1%) were women. Overall, fatigue was a very prevalent long COVID symptom in this cohort (93.5%), with other common (>50%) symptoms being shortness of breath (69.6%), sleeping problems (65.2%), ongoing headaches (64.1%), dizziness (63.0%), heart palpitations (60.9%), brain fog (59.8%), muscular pain (54.3%), and chest tightness (53.3%). Table 1 shows additional descriptives of the 85 participants who had an AS. 36.5% had a BMI in the obesity range (≥30 Kg/m^2^), and 1.2% in the underweight range (<18.5 Kg/m^2^). The majority (62.4%) had third level education and 42.4% were current or former smokers. Other than hypertension (17.6%), prevalences of heart disease and diabetes were very low (<5%), and there were no instances of Parkinson’s disease or other known conditions with risk of autonomic impairment. A fifth were on antidepressant medications and less than 20% were on antihypertensives, beta blockers or benzodiazepines. In terms of COVID-19 history, all but 2 participants were at least 3 months from the onset of COVID-19, a quarter had been hospitalised, and 81.2% met criteria for WHO clinical case definition of post COVID-19 condition. Median scores for CFQ, CES-D and IES-R were 26, 21 and 26, respectively.

**Table 1.** Clinical characteristics of the overall cohort, as well as comparison between OI_AS_ and non-OI_AS_ subgroups. OI_AS_: orthostatic intolerance during active stand; SD: standard deviation; BMI: body mass index; IQR: interquartile range; CFQ: Chalder Fatigue Scale; CES-D: Center for Epidemiological Studies Depression scale; IES-R: Impact of Event Scale – Revised; iOH: initial orthostatic hypotension; cOH_AS_: classical orthostatic hypotension during active stand.

| Characteristic | Overall cohort (n=85) | No OI<sub>AS</sub> (n=29) | OI<sub>AS</sub> (n=56) | P |
| --- | --- | --- | --- | --- |
| Mean age, years (SD) | 46.0 (10.2)<br>(range 25-78) | 49.1 (11.9) | 44.5 (9.0) | 0.075<sup>a</sup> |
| Female sex (%) | 74.1 | 58.6 | 82.1 | 0.019<sup>b*</sup> |
| Mean BMI, Kg/m<sup>2</sup> (SD) | 28.3 (5.1) | 27.2 (4.3) | 28.9 (5.4) | 0.148<sup>a</sup> |
| Mean 5-chair stands time, seconds (SD) | 15.0 (10.4) | 12.8 (5.1) | 16.4 (12.4) | 0.409<sup>a</sup> |
| Third level education (%) | 62.4 | 53.8 | 69.6 | 0.164<sup>b</sup> |
| Previous or current smoker (%) | 42.4 | 56.0 | 40.0 | 0.182<sup>b</sup> |
| History of hypertension (%) | 17.6 | 20.7 | 16.1 | 0.596<sup>b</sup> |
| History of heart disease (%) | 3.5 | 6.9 | 1.8 | 0.267<sup>c</sup> |
| History of diabetes (%) | 3.5 | 6.9 | 1.8 | 0.267<sup>c</sup> |
| On antihypertensive (%) | 16.5 | 24.1 | 12.5 | 0.220<sup>c</sup> |
| On beta blocker (%) | 15.3 | 13.8 | 16.1 | 1.000<sup>c</sup> |
| On antidepressant (%) | 20.0 | 17.2 | 21.4 | 0.647<sup>b</sup> |
| On benzodiazepine (%) | 3.5 | 3.4 | 3.6 | 1.000<sup>c</sup> |
| Median days post-COVID-19 diagnosis (IQR) | 302.0 (333.0)<br>(range 39 – 655) | 249.0 (353.5) | 317.5 (297.8) | 0.628<sup>a</sup> |
| Hospitalised with COVID-19 (%) | 25.9 | 26.9 | 27.8 | 0.936<sup>b</sup> |
| At least 3 months (>91 days) from the onset of COVID-19 (%) | 97.6 | 96.0 | 98.1 | 0.547<sup>c</sup> |
| Post COVID-19 symptoms for at least 2 months (%) | 98.8 | 100 | 98.1 | 1.000<sup>c</sup> |
| In the past month, I have had too little energy to do the things I wanted to do (%) | 83.5 | 76.0 | 96.3 | 0.011<sup>c</sup> |
| Median CFQ score (IQR) | 26.0 (8.0) | 24.0 (10.0) | 27.0 (7.8) | 0.042<sup>a*</sup> |
| Median CES-D score (IQR) | 21.0 (17.0) | 16.0 (16.8) | 24.0 (16.0) | 0.021<sup>a*</sup> |
| Median IES-R score (IQR) | 25.5 (28.8) | 18.5 (30.3) | 31.0 (28.5) | 0.106<sup>a</sup> |
| IOH | 25.9 | 33.3 | 34.1 | 0.952<sup>b</sup> |
| cOH<sub>AS</sub> | 5.9 | 3.6 | 7.5 | 0.654<sup>c</sup> |

<sup>a</sup> 2-sided Mann-Whitney U test; <sup>b</sup> Chi-square test; <sup>c</sup> 2-sided Fisher's exact test; \* statistically significant (P<0.05).

During the AS, 56 participants (65.9%) reported OI_AS_. The frequencies of OI_AS_ symptoms were as follows: “slightly light-headed” (n=31, 55.4%), “light-headed” (n=15, 26.8%), “dizzy” (n=5, 8.9%), “slightly dizzy” (n=4, 7.1%), and “very light-headed” (n=1, 1.8%). 2 of the 85 participants had an early AS termination due to non-hypotensive/cardiac OI_AS_ symptoms (n=1 before the 1^st^ minute, and n=1 before the 3^rd^ minute). Table 1 shows the comparison between OI_AS_ and non-OI_AS_ subgroups. 22 (25.9%) participants fulfilled criteria for iOH, and 5 (5.9%) for cOH_AS_, and neither of the two (P=0.952 an P=0.654, respectively) were significantly associated with OI_AS_. OI_AS_ was more likely in women (P=0.019) and was associated with higher CFQ (P=0.042) and CES-D (P=0.021) scores. The presence of OI_AS_ was more likely to be associated with the activities of daily living impairment criterion used for our application of the WHO clinical case definition (P=0.011), with virtually all participants with OI_AS_ (96.3%) reporting too little energy to do the things they wanted to do in the past month.

In terms of the haemodynamic comparison between OI_AS_ and non-OI_AS_ subgroups (Table 2), participants reporting OI_AS_ had a higher HR at the lowest SBP point before the first minute post-stand (mean HR_nadir_: 88 vs 75 bpm, P=0.004). There were no baseline or subsequent HR differences, or any BP or NIRS differences. In the haemodynamic visualisation in Figure 1, participants’ finishing BP levels (at 3 minutes) seemed higher than at baseline, with 95% CIs around means that clearly did not overlap in the case of DBP (panel b), but without any suggested differences between OI_AS_ and non-OI_AS_ subgroups. On closer inspection, for the overall cohort, there was a statistically significant difference between baseline and 3-minute DBP (mean 81.0 vs 93.1 mmHg, paired samples t-test P<0.001).

**Table 2.**
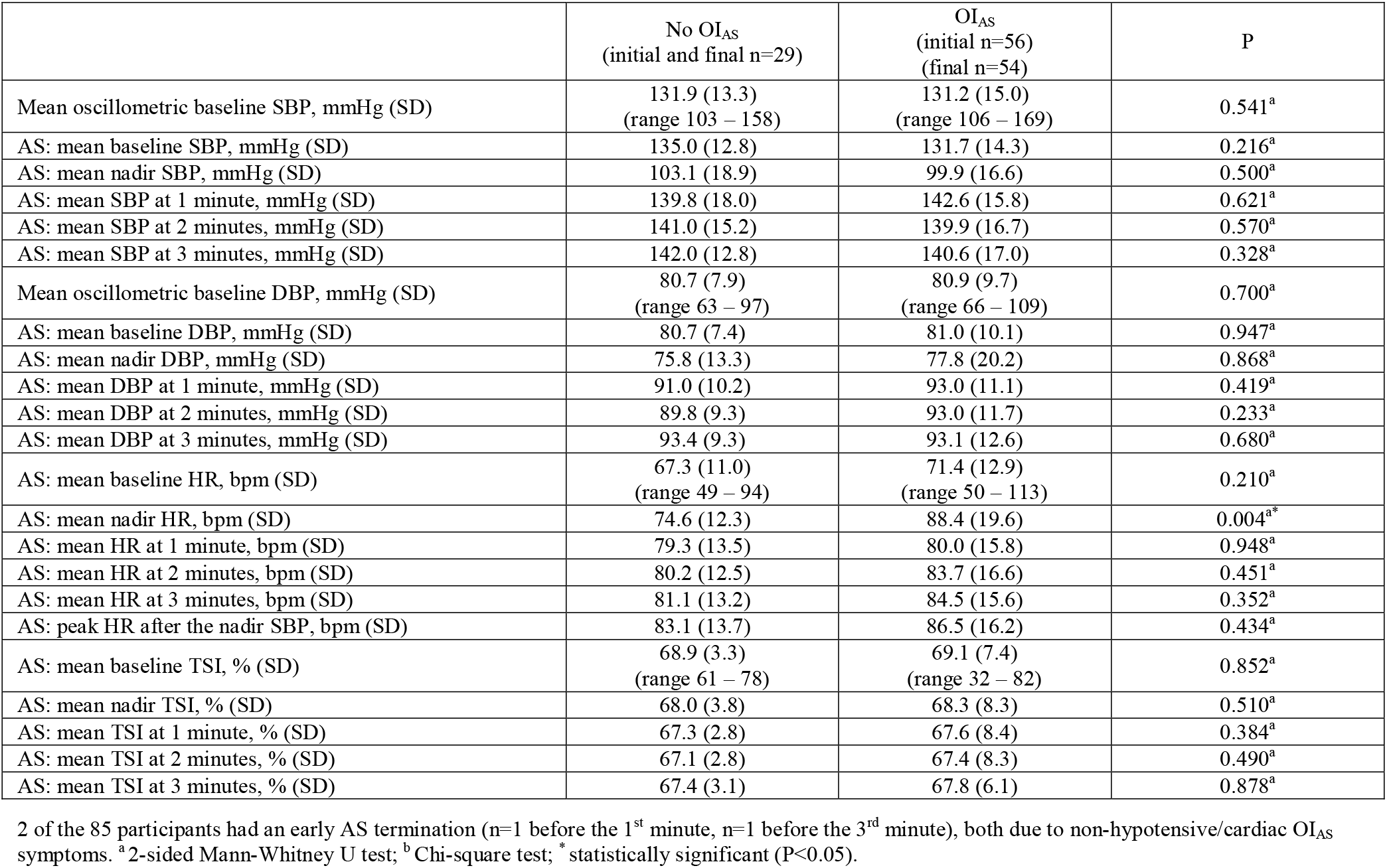
Haemodynamic comparison between OI_AS_ and non-OI_AS_ subgroups. AS: active stand; OI_AS_: orthostatic intolerance during AS; SD: standard deviation; SBP: systolic blood pressure; DBP: diastolic blood pressure; HR: heart rate; bpm: beats per minute; TSI: tissue saturation index.

**Fig. 1.**
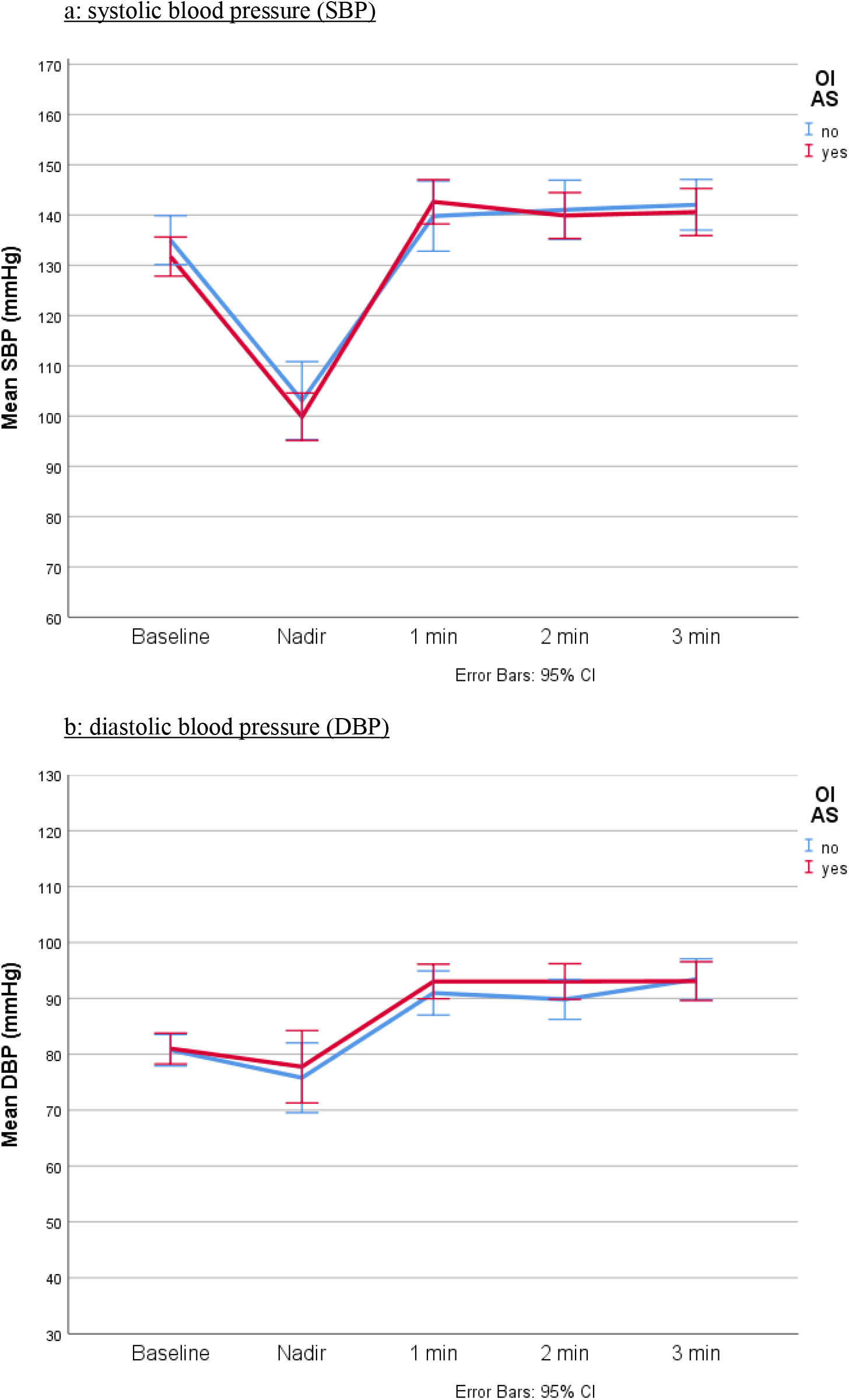
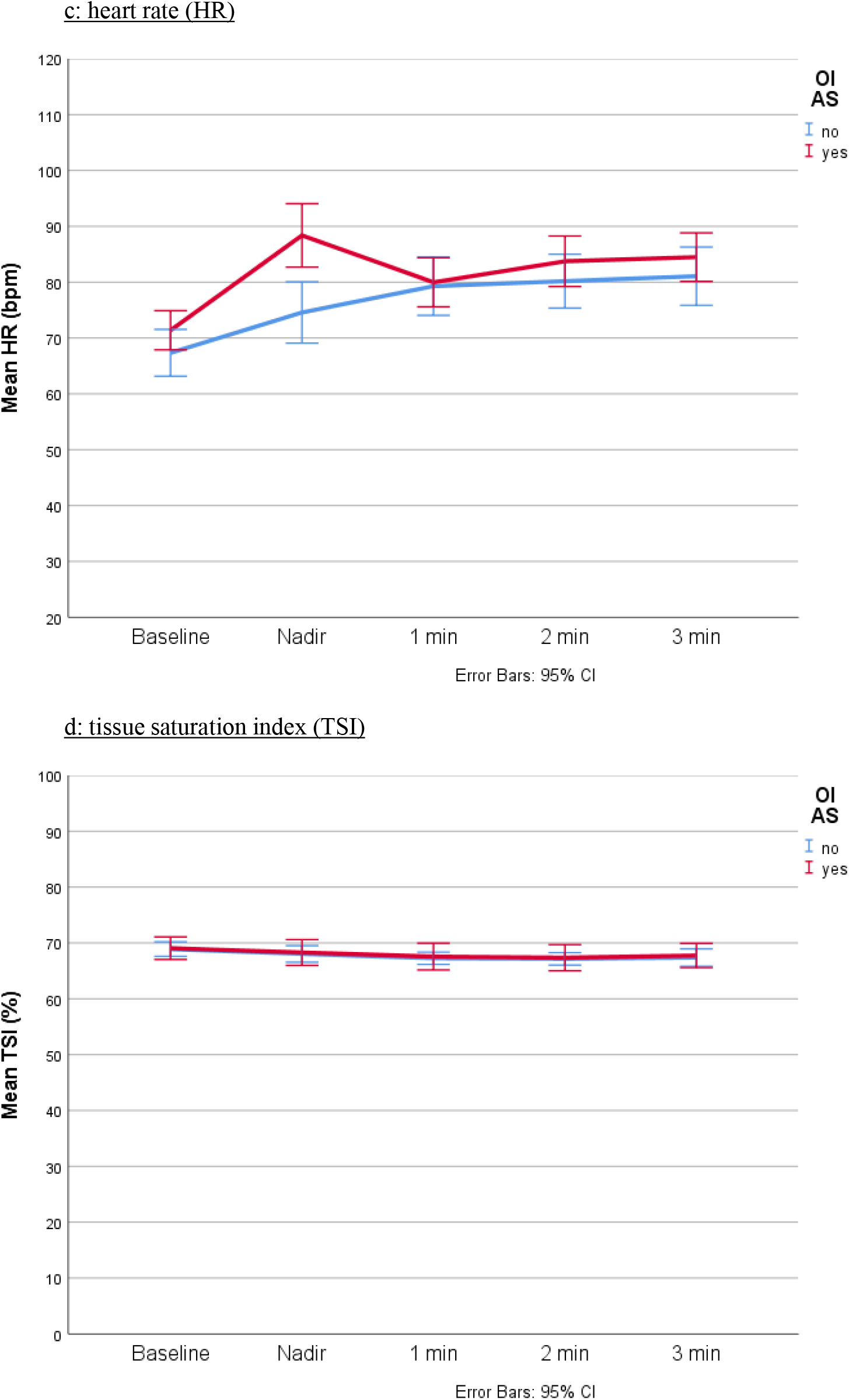
Haemodynamic visualisation of OI_AS_ (n=56) and non-OI_AS_ (n=29) groups; a: systolic blood pressure (SBP); b: diastolic blood pressure (DBP); c: heart rate (HR); d: tissue saturation index (TSI); bpm: beats per minute; CI: confidence interval.

In the logistic regression model to investigate predictors of OI_AS_ (Table 3), the only significant predictor after controlling for age, sex, fatigue, depression, ADL inability, and peak HR after the nadir SBP, HR_nadir_ was the only OI_AS_ predictor (OR=1.09, 95% CI: 1.01-1.18, P=0.027).

**Table 3.** Logistic regression model with predictors of OI_AS_. AS: active stand; CFQ: Chalder Fatigue Scale; CES-D: Center for Epidemiological Studies Depression scale; HR: heart rate; SBP: systolic blood pressure; OR: odds ratio; CI: confidence interval.

|  | OR | 95% CI for OR |  | P |
| --- | --- | --- | --- | --- |
|  |  | Lower | Upper |  |
| Age | 0.98 | 0.91 | 1.05 | 0.604 |
| Female sex | 1.97 | 0.32 | 12.06 | 0.463 |
| CFQ score | 0.96 | 0.80 | 1.16 | 0.666 |
| CES-D score | 1.07 | 0.99 | 1.16 | 0.080 |
| In the past month, I have had too little energy to do the things I wanted to do | 4.48 | 0.32 | 62.08 | 0.263 |
| HR at nadir | 1.09 | 1.01 | 1.18 | 0.027 |
| Peak HR after the nadir SBP | 0.97 | 0.89 | 1.06 | 0.495 |

Of the 85 participants who had an AS, 71 (83.5%) had a tilt table test. 14 participants did not have a tilt for reasons including history of recurrent vasovagal syncope (at least two lifetime episodes), a body weight >120 Kg (tilt table safety limit) or not consenting. All tilt participants had had an AS test. Of them, 28 (39.4%) had OI during tilt (OI_tilt_). The frequencies of OI_tilt_ symptoms were as follows: “slightly light-headed” (n=10, 35.7%), “light-headed” (n=8, 28.6%), “slightly dizzy” (n=3, 10.7%), “dizzy” (n=2, 7.1%), “very light-headed” (n=1, 3.6%), “head spinning” (n=1, 3.6%), “drained” (n=1, 3.6%), “weak” 1 (n=1, 3.6%), and “palpitations” (n=1, 3.6%). No instances of arrhythmia or acute myocardial ischemia were detected in the continuous ECG trace. As regards OI agreement between AS and tilt, 78.6% (n=22) of those who had OI_tilt_ had previously reported OI_AS_ (P=0.020). 18 of the 71 participants had an early tilt termination due to symptoms (n=2 before the 2^nd^ minute, n=3 before the 3^rd^ minute, n=1 before the 4^th^ minute, n=2 before the 5^th^ minute, n=5 before the 6^th^ minute, n=2 before the 8^th^ minute, and n=3 before the 10^th^ minute). Of all the early terminations, 13 (72.2%) were terminated because of OI_tilt_ symptoms (P=0.001). The other 5 early tilt terminations were due to the development of “slight shortness of breath” (n=1), “feet pain” (n=1) and for reasons not related to development of symptoms (n=3). No pre-syncope or syncope occurred in any of the participants. All OI_tilt_ symptoms were reported as transient.

Section 2 in the Supplementary Information shows the comparison between OI_tilt_ and non-OI_tilt_ subgroups. 22 participants (33.3% among the 66 with a tilt of at least 3 minutes) fulfilled criteria for cOH_tilt_, and 7 (13.2% among the 53 with a full 10-minute tilt) for POTS, and neither of the two (P=0.916 an P=0.233, respectively) were significantly associated with OI_tilt_. Among the 7 POTS participants, 2 had a maximum head-up tilt HR of ≥120 beats/minute, 6 had no OI_tilt_, 4 had reported OI_AS_, 2 were on beta blockers, and 1 on antihypertensives. In the 18 participants whose tilt was terminated early, none of the available data fulfilled POTS criteria. As shown in the Supplementary Information, there were no statistically significant differences between OI_tilt_ subgroups across other clinical (section 2) or haemodynamic characteristics (sections 3 and 4), and no significant predictors of OI_tilt_ in the regression model (section 5).

## 4 Discussion

In this study, we investigated predictors of orthostatic intolerance (OI) in adults with long COVID. OI during active stand (OI_AS_) was reported by 66% of our sample and seemed associated with female sex, more fatigue and depressive symptoms, and greater inability to perform activities of daily living, as well as a higher heart rate at the lowest systolic blood pressure point before the first minute post-stand (HR_nadir_). In a regression model also including age, sex, fatigue, depression, ADL inability, and peak HR after the nadir SBP, HR_nadir_ was the only OI_AS_ predictor. 26% of participants had initial and 6% classical orthostatic hypotension, but neither correlated with OI_AS_. Of the participants who had a tilt, 39% had OI during tilt; and of the participants who completed a 10-minute tilt, 13% fulfilled hemodynamic POTS criteria, but most cases (6 out of 7) were asymptomatic.

The HR at the time of nadir SBP after stand seemed more important than the peak HR after the nadir SBP as a predictor of OI_AS_. In this light, findings might reflect different baroreceptor-related HR responses in participants with OI_AS_, possibly due to lower efferent vagus nerve activity, and/or higher sympathetic activation. In this regard, it has been described that with incomplete loss of baroreflex afferents, a mild syndrome of orthostatic tachycardia or orthostatic intolerance may appear; in some cases, it may primarily reflect interruption of efferent right vagus nerve activity, leading to a loss of parasympathetic input to the sinus node, with a consequent increase in heart rate; and in other cases, mild sympathetic activation may occur with stress and provoke tachycardia disproportionate to the increase in blood pressure (Ketch et al., 2002). Indeed, other authors have described the possibility of depressed vagal tone (Leitzke et al., 2020) with or without baroreceptor dysfunction that may lead to tachycardia and heightened cardiac contractility, vascular resistance and venous return (Becker, 2020).

While SARS□CoV□2 might be able to affect neurovascular integrity via direct cytotoxic or indirect pro-inflammatory mechanisms (Khosravani, 2021), our results are in the context of a high burden of psychological symptoms, which is in keeping with other reports (Bucciarelli et al., 2021;Qi et al., 2021). Given our recruitment focus, the proportion of fatigue in our cohort was higher than elsewhere (Akbarialiabad et al., 2021;Lopez-Leon et al., 2021;Sanchez-Ramirez et al., 2021;Sandler et al., 2021). For contextualization to our cohort, previous research showed that a CFQ score of 29 discriminated between chronic fatigue sufferers and controls in 96% of cases [36]; CES-D scores of 16 or greater can signal risk for clinical depression (Lewinsohn et al., 1997); and an IES-R score of 33 and above is suggestive of PTSD (Creamer et al., 2003). Even though in our regression model the HR_nadir_ finding seemed to eclipse previously significant univariate associations with depression and fatigue/ADL inability, adverse psychological states may influence the behaviour of the autonomic nervous system (Peckerman et al., 2003;De Vos et al., 2017); furthermore, in susceptible individuals, discrepancies between predicted and experienced interoceptive signals may engender anxiety during an acute physiological arousal (such as an active stand), which may manifest as transient tachycardia (Miglis and Muppidi, 2017;Owens et al., 2018).

In our cohort there seemed to be evidence of diastolic orthostatic *hyper*tension, fulfilling on average the criterion of a rise in DBP ≥10 mmHg within 3 minutes following AS (Jordan et al., 2020). The pathological significance of this finding is not clear and merits further investigation; indeed, orthostatic hypertension has been found in healthy subjects but also associated with higher (including hypertension) (Jordan et al., 2020) and lower (Wijkman et al., 2016) cardiovascular risks, with more research still needed to clarify its mechanisms and impacts (Jordan et al., 2020). Interestingly, even though orthostatic hypertension did not seem related to OI in our cohort, it has been described that some patients with chronic OI develop symptoms despite a hypertensive response to standing, suggesting that the symptoms of chronic OI may somehow be elicited by central responses to the inappropriate tachycardia, even in the absence of any actual reduction in perfusion pressure (Narkiewicz and Somers, 1998).

In a previous study where autonomic testing was conducted a median of 119 days following acute COVID-19 infection, 22% of patients fulfilled the criteria for POTS (Shouman et al., 2021), in contrast with 13% in our sample with a median delay to testing of 302 days. As required for the POTS definition [39], 6 of our 7 hemodynamically identified POTS had chronic symptoms of OI lasting at least 6 months; however, in 6 out of 7 cases they did not have OI_tilt_ during testing, meaning good tolerability to the tilt challenge and potentially a better clinical prognosis. A previous case report showed no improvement in COVID-19-associated POTS symptoms approximately 5.5 months after symptom onset (Miglis et al., 2020); in a case series of 20 patients, it was reported that most (85%) self-reported residual symptoms 6–8 months after COVID-19, although many felt that they had improved (Blitshteyn and Whitelaw, 2021). Three case reports have documented improvement in POTS after COVID-19 infection, with or without pharmacological support (Ishibashi et al., 2021;O’Sullivan et al., 2021;Ocher et al., 2021). To build on the anecdotal evidence, longitudinal studies are required to assess the evolution of post-COVID POTS in the same cohorts.

Our study has limitations. Firstly, from a study design perspective, generalisability of the findings cannot be assumed given the non-probabilistic recruitment. The evidence we presented is cross-sectional and observational, hence causation cannot be inferred. In addition, we did not have a sample of controls, which can be beneficial in the study of long COVID (Amin-Chowdhury and Ladhani, 2021). Statistical underpower is likely, given the many instances where the statistic of choice for comparisons was the Fisher’s exact test, and a small sample size that precluded inclusion of a greater number of predictors in the regression models.

Another limitation is that our testing protocol did not include other standardised autonomic tests such as heart rate variability with paced breathing or blood pressure response to Valsalva maneuver. However, in the same clinical environment, Townsend *et al*. performed those tests on a different long COVID cohort and reported negative findings (Townsend et al., 2021). Our study did not have more detailed measures of baroreflex function, or any imaging or biomarker information (e.g., hematological, biochemical, immunological). For ethical approval reasons, in some cases our research tilts had to be stopped sooner (e.g., with only mild symptoms) than is often the case for tilts used in clinical practice to look for full symptom reproduction. From a haemodynamic data processing point of view, other studies have extracted the raw data from the Finapres® and performed signal averaging prior to analyses, for example in 5-second bins (van der Velde et al., 2007). While *post-hoc* signal averaging can theoretically reduce the risk of spurious observations due to signal artifacts (Finucane et al., 2019), in this study we followed the direct observation method that is routinely utilised in clinical practice for the contemporaneous clinical assessment of patients.

In conclusion, in our study OI_AS_ was associated with a higher initial HR on AS, which after 1 minute equalized with the non-OI_AS_ group, and POTS was infrequent and largely asymptomatic. The burden of psychological symptoms in this cohort was high and findings may be related to interoceptive mechanisms. However, more research is required to understand the mechanisms and long-term prognosis of autonomic function in long COVID, to better delineate therapies and estimate the need for services.

## Data Availability

The datasets generated and analyzed for this study are not publicly available due to ethical approval reasons.

## 6 Author Contributions

Conceptualization: Roman Romero-Ortuno, Ann Monaghan; Methodology: Roman Romero-Ortuno, Ann Monaghan, Lisa Byrne; Clinical data collection: Ann Monaghan, Glenn Jennings, Feng Xue, Eoin Duggan, Roman Romero-Ortuno; Formal analysis and investigation: Roman Romero-Ortuno, Ann Monaghan, Glenn Jennings, Eoin Duggan; Writing – original draft preparation: Roman Romero-Ortuno; Writing – review and editing: Ann Monaghan, Glenn Jennings, Feng Xue, Eoin Duggan, Lisa Byrne; Funding acquisition: Roman Romero-Ortuno; Resources: Roman Romero-Ortuno, Lisa Byrne; Supervision: Eoin Duggan, Lisa Byrne, Roman Romero-Ortuno.

## 8 Acknowledgments

We are very grateful to our participants for their involvement in the study.

## 10 Supplementary Material

Please see the Supplementary Material section for further information.

