## Supplementary Material for "Orthostatic intolerance in adults with long COVID was not associated with postural orthostatic tachycardia syndrome"

### 1 COVID-19 and non-COVID-19 exclusion criteria for enrolment.

COVID-19-related exclusion criteria were: (1) being in the acute COVID-19 phase and/or experiencing any of the following symptoms/situations included in a COVID-19 Health Screening Assessment Tool administered prior to enrolment: (2) new cough or shortness of breath, fever or chills within the last 48 hours; (3) new loss/distortion/alteration of sense of smell or taste; sore throat/swollen glands; new headaches; vomiting; diarrhoea; (4) positive COVID-19 test in the last 14 days; (5) awaiting a test result for COVID-19; (6) in close contact with someone who had COVID-19 symptoms in the last 14 days; (7) visit to a residential institution in the last 14 days where ongoing COVID-19 transmission had been confirmed; (8) being from a geographical region where there was ongoing COVID-19 transmission and on lockdown/travel restriction; (9) international travel in the last 14 days; or (10) any other symptoms/situations as per Principal Investigator’s clinical judgement.

### Non-COVID-19 exclusion criteria were: (11) being pregnant; (12) cognitive impairment/dementia with inability to give informed consent; (13) upper limb lymphoedema (cuff-site); and (14) severe skin allergies to adhesive tapes. The following were contra-indications for tilt testing: (15) stroke or acute myocardial infarction within the past 90 days (or any other acute cardiac event); (16) unstable angina; (17) uncontrolled cardiac arrhythmias causing symptoms or hemodynamic compromise; (18) uncontrolled symptomatic heart failure; (19) symptomatic severe aortic stenosis; (20) suspected or known dissecting aortic aneurysm; (21) acute myocarditis or pericarditis; (22) acute pulmonary embolus or pulmonary infarction; (23) patients in whom low organ perfusion pressures may compromise end-artery supplied tissue; (24) severe left ventricular outflow obstruction; and (25) critical mitral stenosis. In addition, for safety reasons, participants who during the anamnesis reported a history of (26) recurrent vasovagal syncope (at least two lifetime episodes) or had a (27) body weight >120 Kg were excluded from tilt testing.

### 2 Comparison between OI_tilt_ and non-OI_tilt_ subgroups. OI_tilt_: orthostatic intolerance during tilt; OI_AS_: orthostatic intolerance during active stand; SD: standard deviation; BMI: body mass index; IQR: interquartile range; CFQ: Chalder Fatigue Scale; CES-D: Center for Epidemiological Studies Depression scale; IES-R: Impact of Event Scale – Revised; cOH_tilt_: classical orthostatic hypotension during tilt; POTS: postural orthostatic tachycardia syndrome.

| Characteristic | No OI_tilt_ (n=43) | OI_tilt_ (n=28) | P |
| --- | --- | --- | --- |
| Mean age, years (SD) | 47.1 (10.6) | 44.1 (9.7) | 0.281^a^ |
| Female sex (%) | 69.8 | 75.0 | 0.632^b^ |
| Third level education (%) | 65.9 | 63.0 | 0.807^b^ |
| Previous or current smoker (%) | 51.3 | 40.7 | 0.399^b^ |
| History of hypertension (%) | 20.9 | 14.3 | 0.479^b^ |
| History of heart disease (%) | 4.7 | 0.0 | 0.515^c^ |
| History of diabetes (%) | 7.0 | 0.0 | 0.273^c^ |
| Median days post-COVID-19 diagnosis (IQR) | 269.0 (327.0) | 234.0 (339.5) | 0.767^a^ |
| Hospitalised with COVID-19 (%) | 26.8 | 26.9 | 0.993^b^ |
| Median CFQ score (IQR) | 25.5 (9.8) | 25.0 (8.5) | 0.994^a^ |
| Median CES-D score (IQR) | 19.0 (16.5) | 23.0 (19.0) | 0.267^a^ |
| Median IES-R score (IQR) | 22.0 (31.0) | 26.0 (31.0) | 0.585^a^ |
| Mean BMI, kg/m^2^ (SD) | 28.0 (5.4) | 28.6 (4.9) | 0.410^a^ |
| Mean 5-chair stands time, seconds (SD) | 14.8 (10.5) | 13.8 (9.3) | 0.827^a^ |
| OI_AS_ | 51.2 | 78.6 | 0.020^b^ |
| cOH_tilt_ | 31.0 | 32.1 | 0.916^b^ |
| POTS | 14.3 | 3.7 | 0.233^c^ |
| On antihypertensive (%) | 18.6 | 14.3 | 0.753^c^ |
| On beta blocker (%) | 9.3 | 21.4 | 0.177^c^ |
| On antidepressant (%) | 11.6 | 28.6 | 0.071^b^ |
| On benzodiazepine (%) | 2.3 | 3.6 | 0.100^c^ |

^a^ 2-sided Mann-Whitney U test; ^b^ Chi-square test; ^c^ 2-sided Fisher’s exact test; ^*^ statistically significant (P<0.05)

### 3 Haemodynamic comparison between OItilt and non-OItilt subgroups. OItilt: orthostatic intolerance during tilt; SD: standard deviation; SBP: systolic blood pressure; DBP: diastolic blood pressure; HR: heart rate; bpm: beats per minute; TSI: tissue saturation index.

|  | No OI_tilt_  (initial n=43)  (final n=38) | OI_tilt_  (initial n=28)  (final n=15) | P |
| --- | --- | --- | --- |
| Mean oscillometric baseline SBP, mmHg (SD) | 133.0 (11.7)  (range 112 – 160) | 134.9 (13.7)  (range 105 – 162) | 0.504^a^ |
| Tilt: mean baseline SBP, mmHg (SD) | 130.1 (13.7) | 130.7 (11.3) | 0.801^a^ |
| Tilt: mean nadir SBP, mmHg (SD) | 123.1 (17.3) | 121.2 (22.0) | 0.666^a^ |
| Tilt: mean SBP at 1 minute, mmHg (SD) | 131.6 (15.9) | 132.3 (17.0) | 0.970^a^ |
| Tilt: mean SBP at 2 minutes, mmHg (SD) | 132.3 (19.6) | 133.1 (15.9) | 0.707^a^ |
| Tilt: mean SBP at 3 minutes, mmHg (SD) | 133.0 (18.0) | 133.4 (19.6) | 0.989^a^ |
| Tilt: mean SBP at 4 minutes, mmHg (SD) | 135.7 (20.0) | 133.5 (19.9) | 0.933^a^ |
| Tilt: mean SBP at 5 minutes, mmHg (SD) | 132.2 (17.8) | 134.4 (14.5) | 0.538^a^ |
| Tilt: mean SBP at 6 minutes, mmHg (SD) | 132.6 (22.0) | 131.0 (15.6) | 0.699^a^ |
| Tilt: mean SBP at 7 minutes, mmHg (SD) | 134.6 (20.8) | 130.2 (16.8) | 0.583^a^ |
| Tilt: mean SBP at 8 minutes, mmHg (SD) | 132.1 (16.9) | 132.9 (14.7) | 0.682^a^ |
| Tilt: mean SBP at 9 minutes, mmHg (SD) | 134.3 (16.2) | 132.3 (16.2) | 0.585^a^ |
| Tilt: mean SBP at 10 minutes, mmHg (SD) | 137.9 (17.9) | 128.5 (15.9) | 0.069^a^ |
| Mean oscillometric baseline DBP, mmHg (SD) | 81.2 (9.4)  (range 66 – 104) | 83.9 (9.8)  (range 64 – 99) | 0.169^a^ |
| Tilt: mean baseline DBP, mmHg (SD) | 78.3 (10.5) | 79.4 (9.9) | 0.449^a^ |
| Tilt: mean nadir DBP, mmHg (SD) | 81.6 (14.3) | 82.0 (17.9) | 0.635^a^ |
| Tilt: mean DBP at 1 minute, mmHg (SD) | 86.0 (11.0) | 89.6 (16.3) | 0.494^a^ |
| Tilt: mean DBP at 2 minutes, mmHg (SD) | 88.1 (14.4) | 92.6 (16.9) | 0.347^a^ |
| Tilt: mean DBP at 3 minutes, mmHg (SD) | 87.0 (12.4) | 88.2 (19.1) | 0.479^a^ |
| Tilt: mean DBP at 4 minutes, mmHg (SD) | 89.0 (14.9) | 88.6 (17.9) | 0.989^a^ |
| Tilt: mean DBP at 5 minutes, mmHg (SD) | 86.6 (13.9) | 88.8 (13.6) | 0.816^a^ |
| Tilt: mean DBP at 6 minutes, mmHg (SD) | 88.1 (14.4) | 90.8 (14.4) | 0.705^a^ |
| Tilt: mean DBP at 7 minutes, mmHg (SD) | 88.5 (15.4) | 91.9 (15.4) | 0.536^a^ |
| Tilt: mean DBP at 8 minutes, mmHg (SD) | 89.0 (12.3) | 90.5 (16.6) | 0.913^a^ |
| Tilt: mean DBP at 9 minutes, mmHg (SD) | 88.9 (13.1) | 91.6 (13.7) | 0.655^a^ |
| Tilt: mean DBP at 10 minutes, mmHg (SD) | 90.9 (15.1) | 87.5 (14.3) | 0.337^a^ |
| Tilt: mean baseline HR, bpm (SD) | 67.4 (10.0)  (range 47 – 91) | 64.7 (12.3)  (range 46 – 95) | 0.166^a^ |
| Tilt: mean nadir HR, bpm (SD) | 74.7 (15.2) | 71.8 (17.0) | 0.193^a^ |
| Tilt: mean HR at 1 minute, bpm (SD) | 77.6 (14.5) | 76.8 (18.4) | 0.341^a^ |
| Tilt: mean HR at 2 minutes, bpm (SD) | 78.2 (14.3) | 77.8 (14.1) | 0.812^a^ |
| Tilt: mean HR at 3 minutes, bpm (SD) | 78.5 (14.6) | 80.5 (18.4) | 0.934^a^ |
| Tilt: mean HR at 4 minutes, bpm (SD) | 78.6 (13.4) | 78.7 (14.3) | 0.933^a^ |
| Tilt: mean HR at 5 minutes, bpm (SD) | 80.7 (13.6) | 80.1 (11.6) | 0.942^a^ |
| Tilt: mean HR at 6 minutes, bpm (SD) | 80.3 (13.3) | 81.2 (19.0) | 0.738^a^ |
| Tilt: mean HR at 7 minutes, bpm (SD) | 80.6 (14.1) | 83.2 (17.8) | 0.925^a^ |
| Tilt: mean HR at 8 minutes, bpm (SD) | 80.0 (14.2) | 80.5 (19.6) | 0.548^a^ |
| Tilt: mean HR at 9 minutes, bpm (SD) | 81.7 (14.9) | 81.2 (18.2) | 0.428^a^ |
| Tilt: mean HR at 10 minutes, bpm (SD) | 80.3 (14.8) | 82.0 (23.0) | 0.724^a^ |
| Tilt: mean baseline TSI, % (SD) | 70.0 (4.7)  (range 56 – 79) | 69.5 (4.5)  (range 62 – 84) | 0.450^a^ |
| Tilt: mean nadir TSI, % (SD) | 69.1 (5.1) | 69.1 (3.8) | 0.793^a^ |
| Tilt: mean TSI at 1 minute, % (SD) | 68.9 (4.5) | 68.4 (3.7) | 0.521^a^ |
| Tilt: mean TSI at 2 minutes, % (SD) | 67.9 (5.0) | 67.8 (3.3) | 0.677^a^ |
| Tilt: mean TSI at 3 minutes, % (SD) | 67.9 (5.8) | 68.2 (3.4) | 0.868^a^ |
| Tilt: mean TSI at 4 minutes, % (SD) | 68.1 (6.0) | 68.0 (3.5) | 0.577^a^ |
| Tilt: mean TSI at 5 minutes, % (SD) | 68.0 (6.0) | 67.8 (4.1) | 0.515^a^ |
| Tilt: mean TSI at 6 minutes, % (SD) | 68.4 (5.5) | 66.6 (3.1) | 0.091^a^ |
| Tilt: mean TSI at 7 minutes, % (SD) | 68.5 (5.4) | 67.2 (2.9) | 0.155^a^ |
| Tilt: mean TSI at 8 minutes, % (SD) | 69.0 (5.6) | 67.5 (3.2) | 0.132^a^ |
| Tilt: mean TSI at 9 minutes, % (SD) | 68.7 (5.3) | 67.7 (3.1) | 0.142^a^ |
| Tilt: mean TSI at 10 minutes, % (SD) | 69.3 (4.0) | 69.7 (5.9) | 0.892^a^ |

18 of the 71 participants had an early tilt termination (n=2 before the 2^nd^ minute, n=3 before the 3^rd^ minute, n=1 before the 4^th^ minute, n=2 before the 5^th^ minute, n=5 before the 6^th^ minute, n=2 before the 8^th^ minute, and n=3 before the 10^th^ minute). Of all the early terminations, 13 (72.2%) were terminated because of OI_tilt_ symptoms (P=0.001). The other 5 early tilt terminations were due to the development of “slight shortness of breath” (n=1), “feet pain” (n=1) and for reasons not related to symptom development (n=3). ^a^ 2-sided Mann-Whitney U test; ^b^ Chi-square test; ^*^ statistically significant (P<0.05).

### 4 Haemodynamic visualisation of OItilt and non-OItilt groups: a: systolic blood pressure (SBP); b: diastolic blood pressure (DBP); c: heart rate (HR); d: tissue saturation index (TSI). CI: confidence interval.

a: systolic blood pressure (SBP)

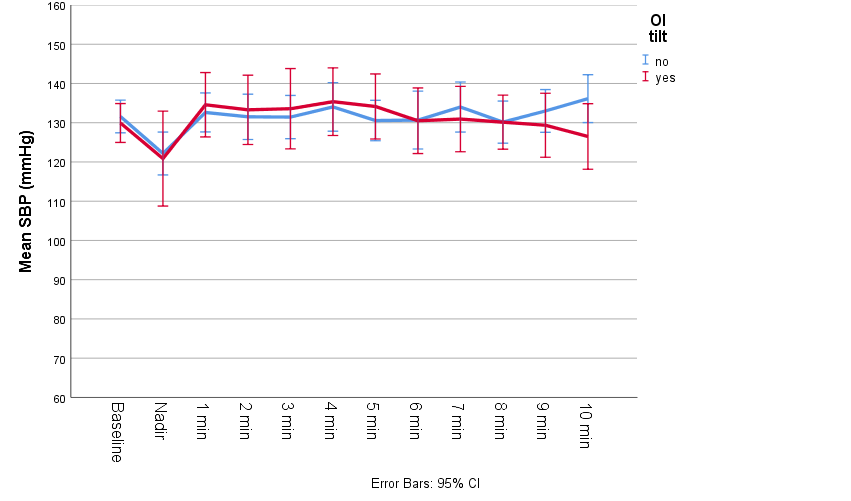

b: diastolic blood pressure (DBP)

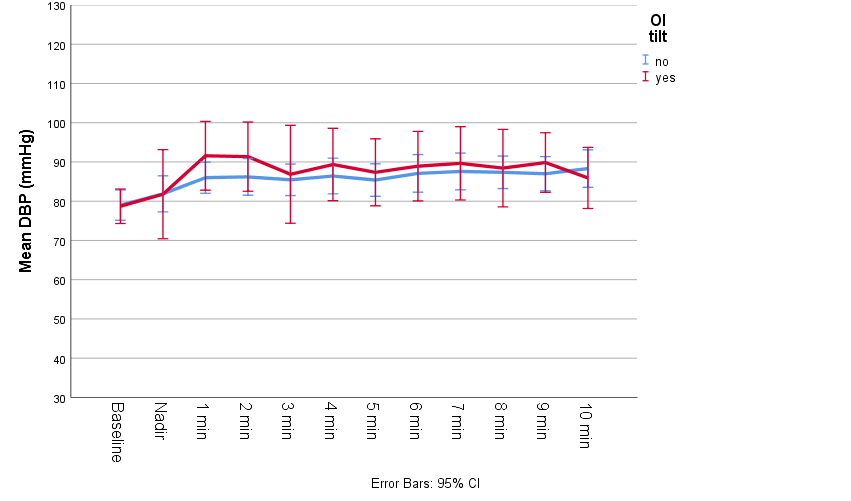

c: heart rate (HR)

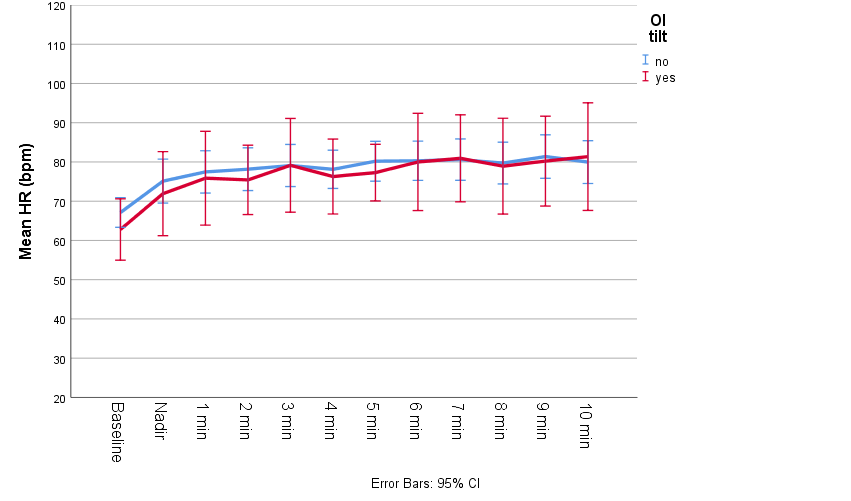

d: tissue saturation index (TSI)

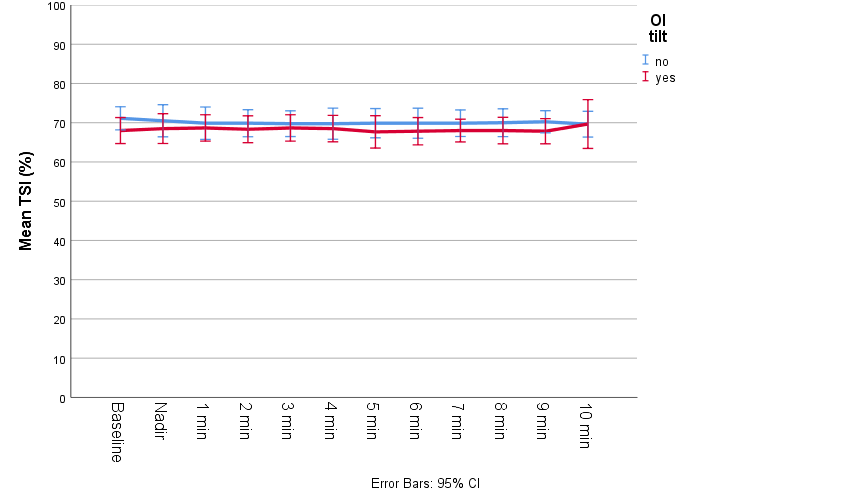

### 5 Logistic regression model with predictors of OItilt. OItilt: orthostatic intolerance during tilt; CFQ: Chalder Fatigue Scale; CES-D: Center for Epidemiological Studies Depression scale; cOHtilt: classic orthostatic hypotension during tilt; POTS: postural orthostatic tachycardia syndrome; OR: odds ratio; CI: confidence interval.

|  | OR | 95% C.I. for OR | | P |
| --- | --- | --- | --- | --- |
|  |  | Lower | Upper |  |
| Age | 0.96 | 0.90 | 1.02 | 0.202 |
| Female sex | 2.99 | 0.71 | 12.65 | 0.136 |
| CFQ score | 0.97 | 0.86 | 1.08 | 0.547 |
| CES-D score | 1.02 | 0.97 | 1.08 | 0.435 |
| cOH_tilt_ | 1.04 | 0.28 | 3.79 | 0.955 |
| POTS | 0.15 | 0.01 | 1.53 | 0.109 |
| Lowest SBP after tilt | 1.00 | 0.97 | 1.04 | 0.858 |
